# The ocrelizumab phase II extension trial suggests the potential to improve the risk:benefit balance in multiple sclerosis

**DOI:** 10.1101/2020.01.09.20016774

**Authors:** David Baker, Gareth Pryce, Louisa K. James, Monica Marta, Klaus Schmierer

## Abstract

**Objective:** Ocrelizumab inhibits relapsing multiple sclerosis when administered every six months. Based on potential similar memory B cell depletion mechanisms with cladribine and alemtuzumab, we hypothesised that CD20-depletion of B cells by ocrelizumab may exhibit a duration of response exceeding the current licenced treatment interval.

**Methods:** Internet-located information from regulatory submissions and meeting reports relating to the unpublished open-label, phase II ocrelizumab extension trial (NCT00676715) were reviewed. This followed people (54-55/arm) with MS, who switched from placebo or interferon-beta to ocrelizumab for three 600mg treatment cycles (week 24, 48, 72) or people treated with ocrelizumab for four 600mg treatment cycles (week 0-72), followed by an 18 month treatment-free period.

**Results:** CD19+ B cells were rapidly depleted within 2 weeks and slow CD19+ B cell repopulation began about 6 months after the last infusion with median-repletion of over 15 months. The reduced annualized relapse rate during the published efficacy study appeared to be maintained in the extension study and there were no new T1 gadolinium-enhancing or T2 lesions detected in the treatment-free period. Importantly, within these extension cohorts, there appeared to be fewer adverse events and infections events.

**Conclusions:** Ocrelizumab appears to induce durable relapsing disease inhibition, within 3 treatment cycles Therefore, it may be possible to reduce the frequency of dosing to maintain efficacy, whilst limiting infection and other risks associated with continuous immunosuppression. Further studies are now clearly required to determine whether this data is robust, as few people seemed to complete the study.

## INTRODUCTION

Multiple sclerosis (MS) is the major demyelinating disease of the central nervous system. Although considered to be a T cell-mediated disease, CD20 B cell–depleting antibodies exhibit high efficacy in MS (1-3). Indeed, we have suggested that agents that inhibit relapsing MS all target memory B cell populations (4-6). These may act directly on B cells or may target T cells directly of secondary through loss of B cell help for T cells (1, 4, 7, 8). Although the B cell subset depletion potential of ocrelizumab has yet to be fully reported, it and rituximab potently deplete memory B cells (1,7,9). Efficacy of CD20-depletion develops within a few weeks of treatment-onset and is typically administered in 6 monthly cycles to permanently deplete CD19+ B cell populations, which includes memory B cells (1,3,7,10). However, memory B cell depletion can last for years following treatment, probably due to their slow repopulation kinetics (7, 11). This suggests that there may be durable efficacy beyond 6 months, as suggested from studies with rituximab (1). Furthermore, marked memory B cell depletion appears to be a common mechanism contributing to the efficacy of alemtuzumab and cladribine (5,6). These are considered immune-reconstitution therapies with long-term efficacy from a short-term treatment cycle (12-14), so we hypothesized that ocrelizumab could similarly induce benefit extending beyond a six-monthly treatment cycle.

This is important, because while ocrelizumab use has been well-tolerated in MS (3), B cells form a central part of immunity. As such, continuous B cell depletion is associated with eventual hypogammaglobulinaemia creating an increased risk of infection and reduced vaccination efficacy (14,15). These risks appear to be significant, as the development of ocrelizumab was terminated in other CD20-responsive autoimmunities because of infection-related fatalities adversely affecting the risk: benefit balance (16). It is therefore important to determine whether efficacy can be maintained and complications de-risked by reduced-frequency dosing. This strategy is currently being tested in natalizumab with a view to reduce the risk of developing progressive multifocal leucoencephalopathy (17). Furthermore, preliminary studies with rituximab may suggest that dosing to memory B cell population kinetics can reduce dosing frequency whilst maintaining efficacy (18)

Despite policies to make trial-data available, trial-information presented at major international conferences are not always followed by peer-reviewed publications (5, 19). Therefore, information cannot easily be searched or interrogated by internet engines and fails to become common knowledge and perhaps allows people to be unwittingly exposed to unnecessary safety issues (19). Although the phase II (NCT00676715) and phase III (NCT03599245) ocrelizumab efficacy MS studies are published (2,3), the extension study data remain unpublished, except in abstract form (20-22). Whilst the phase III extension study examined the influence of 6-monthly dosing (22), the phase II extension study followed people during a significant treatment-free period (20,21). This suggests that clinical benefits are maintained after an 18-month treatment-free period, which may have risk:benefit implications.

## METHODOLOGICAL APPROACH

### Data Analysis

Information on the phase II ocrelizumab trial extension studies have been presented from 2012 onwards (20,21). Through meeting abstracts, posters and regulatory documents available on the internet, we were able to determine the key trial results. Data was extracted, with the assistance of WebplotDigitizer V4.1 (https://automeris.io/WebPlotDigitizer. A Rohatgi) and a facsimile of presented data is reported. Attempts to verify these data with Freedom of Information requests submitted to the European Medicines Agency (5) were unsuccessful. Access to a redacted copy of the WA21493 clinical study report March 2016 was rejected by Roche on the grounds that the trial is ongoing until 9 December 2021.

### Trial Design

Details of the phase II placebo-controlled, randomised double-blind trial (NTC00676715) of placebo, beta-interferon-1a (IFNβ) and two doses of ocrelizumab and the demographics and numbers of participants have been published (2,20). In brief, this included the requirement to have had two documented relapses or ≥1 relapse and ≥ 6 T2 lesions within the year prior to screening and to have an EDSS of 1.0-6.0 (2). Likewise, the methods and outcomes of the 24 week blinded trial have been reported (2). In brief, people with MS (pwMS) were randomised to either: (a) two placebo intravenous (i.v.) infusions at 15 day intervals (b) two infusions of 300mg ocrelizumab at 15 day intervals (600mg dose), with infusion reaction prophylaxis, and (c) open-label 30μg IFNβ administered twice a week (2). An additional group of people, not discussed here, received two doses of 1000mg ocrelizumab (2). At 24 weeks all groups (a-c) received ocrelizumab, which was administered at weeks 24, 48 and 72. People treated with placebo and IFNβ initiated their ocrelizumab treatment with two doses of 300mg ocrelizumab; all subsequent doses were single infusions of 600mg at six-month intervals consistent with the subsequent phase III trial (3) and the current labels in the United States and Europe (23,24). These dosing schedules induced comparable levels of T and B cell depletion (25,26). Those pwMS receiving high dose (1000mg) ocrelizumab received a single 1000mg infusion at weeks 24 and 48 and 600mg at week 72 (20,21). To allow for a definitive trial report to be written by the company, only data related to the 600mg licenced dose (23,24) are reported here. The treatment response was monitored to week 96 at which time people entered a treatment-free extension period for examination of a safety follow-up over 24 weeks and a B cell monitoring period up to week 144, where pwMS were assessed every 12 weeks until repletion, defined as a return to the baseline CD19 count or the lower limit of normal, occurred (20). PwMS who discontinued the 96 week treatment period also underwent the treatment–free, safety follow-up period (**Table 1**). Safety and efficacy were assessed throughout the study via regular neurological and physical examination (2,20).

**Table 1.** Durable efficacy of ocrelizumab following a short term treatment cycle

| Treatment | mAb Cycles | Trial | Analysis | Relapse Rate (aARR) |  |
| --- | --- | --- | --- | --- | --- |
| Placebo |  | Phase II | First 24 weeks (0-24) | 0.557 | P=0.0019 |
| Ocrelizumab | 1 cycle | Phase II | First 24 weeks (0-24) | 0.127 |  |
| Ocrelizumab-Interferonβ | 1 cycle | Phase II OLE | First 24 weeks (24-48) | 0.131 | |
| Ocrelizumab-Placebo | 1 cycle | Phase II OLE | First 24 weeks (24-48) | 0.200 |  |
| Ocrelizumab-Interferonβ | 3 cycles | Phase II OLE | Last 24 weeks (120-144) | 0.068 | |
| Ocrelizumab-Placebo | 3 cycles | Phase II OLE | Last 24 weeks (120-144) | 0.035 |  |
| Ocrelizumab | 4 cycles | Phase II OLE | Last 24 weeks (120-144) | 0.075 |  |
| Interferonβ | | Phase III | First 48 weeks (0-48) | 0.274 | P<0.001 |
| Ocrelizumab | 2 cycles | Phase III | First 48 weeks (0-48) | 0.141 |  |
| Ocrelizumab | 2 cycles | Phase III OLE | First 48 weeks (96-144) | 0.098 |  |
| Ocrelizumab-Interferonβ | 6 cycles | Phase III OLE | Last 48 weeks (192-240) | 0.072 | |
| Ocrelizumab | 6 cycles | Phase III OLE | Last 48 weeks (96-144) | 0.103 |  |
| Ocrelizumab-Placebo | 3 cycles | Phase II OLE | Last 48 weeks (96-144) | 0.116 |  |
| Ocrelizumab-Interferonβ | 3 cycles | Phase II OLE | Last 48 weeks (96-144) | 0.076 | |

Data on the reported adjusted annualised relapse rate (aARR) were extracted from meeting reports relating to the phase II and the phase II open label extension (OLE) following treatment with ocrelizumab (Week 0-96. N=51-46), placebo (Week 0-24. N=54) switching to ocrelizumab (week 24-96 N=51-48) or twice weekly interferon beta (week 0-24. N=51) switching to ocrelizumab N=49-46), and the phase III (OPERA I & OPERA II) following treatment with interferon beta N=829 and ocrelizumab (N=827) and the phase III OLE extension study following people treated with ocrelizumab (n = 702) or Beta interferon (Week 96-240. N=702-570). Probability compared ocrelizumab to placebo (phase II) or interferon beta (phase III) studies (2, 20-22)

## RESULTS/FINDINGS

The phase II study consisted of a screening period, where 273 pwMS were assessed for eligibility and 220 were entered into randomisation. Fifty-four pwMS were treated in the placebo arm, 54 pwMS in the IFNβ arm and 55 pwMS in each of two ocrelizumab arms (20). All pwMS in the placebo arm, 94% on IFNβ and 93% on 600mg ocrelizumab infusions completed the phase II efficacy study (20). In these three arms, 151 (93%) pwMS chose to enter the open-label extension study to receive ocrelizumab from week 24-96 and 133 (82%) pwMS subsequently entered the treatment-free follow-up period (Week 96-144), which also acquired data from people who had withdrawn from the study during earlier cycles. The follow-up study from week 96-144 provided time to monitor B cell repopulation. 140 people completed 96 weeks and the safety follow-up to 120 weeks. However, relatively few people (n=33) are reported to have week 144 data, and therefore, caution must be applied when viewing the results (20).

Analysis of lymphocyte subsets indicated that ocrelizumab induced a rapid and marked depletion of CD19+ B cells within two weeks, and maintained a nadir state for the initial 24 week phase II efficacy study and the 96 week extension study following switching of placebo and IFNβ-treated to 600mg infusions of ocrelizumab (**Figure 1A**). Slow CD19+ B cell repopulation began about 6 months after the last infusion (**Figure 1A**). The median time to B cell repletion was 62 weeks (95%CI 60-72 weeks)/placebo arm and 71.9 weeks (95% CI 62.3-75.7 (20,21,25) (**Figure 1B**). This occurred in a median of 70% of pwMS following 3 cycles and 52% of pwMS following 4 cycles ocrelizumab by week 144 (**Figure 1B**). The duration of depletion ranged from 27-175 weeks (23,25). Despite being part of the trial protocol (WA21493), depletion and repopulation of B cell subset data were not publicly available. However, based on studies with rituximab, we would anticipate that repopulation of memory B cells would take significantly longer than apparent CD19 B cell repletion, which is largely driven by repopulation of immature/mature B cells (1,7). A small fraction of people appeared not to deplete adequately in those receiving less than 2 cycles of ocrelizumab (**Figure 1B**). Whether this relates to polymorphisms influencing antibody-dependent cell cytotoxicity remains to be established. Examination of CD4 and CD8 T cell and natural killer cell levels demonstrated very little change although there was a transient drop following the initial infusion of ocrelizumab (**Figure 1C**). This was not really evident when placebo and IFNβ arms were switched, as bloods were only collected 12 weeks after infusion (**Figure 1C**). These are consistent with effects reported in phase III studies (27).

**FIGURE1.**
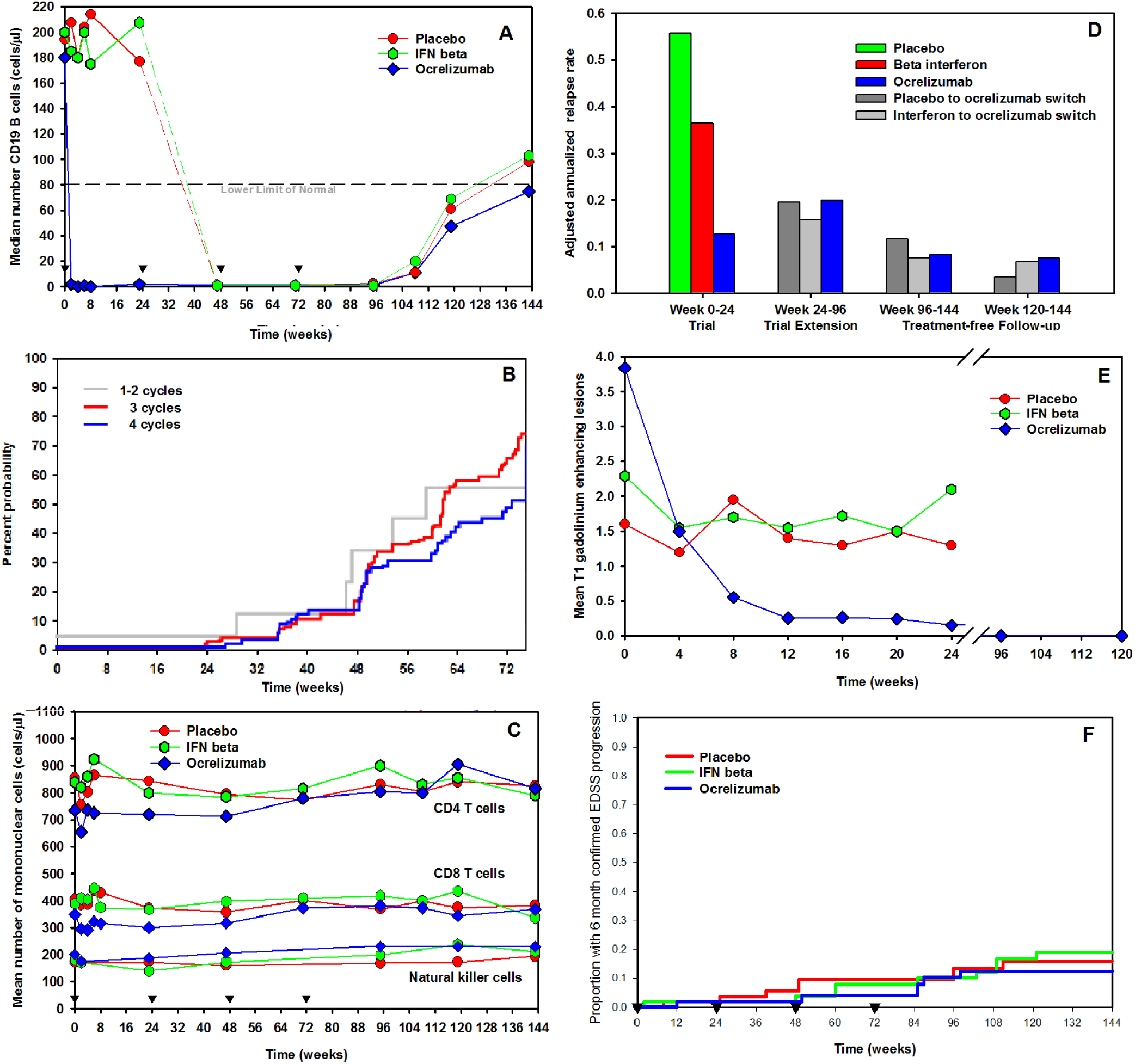
Inhibition of CD19 B lymphocytes by ocrelizumab. PwMS were randomised to 1 cycle (inverse triangles) of i.v. placebo (n=54), i.m. interferon beta 1a (n=54) or 600mg i.v. ocrelizumab (n=55) followed by 3 six-monthly cycles (inverse triangles) of 600mg ocrelizumab and a treatment-free period from week 72. (A) The median absolute number of peripheral blood CD19+ B cells was assessed by flow cytometry (B) The median time taken to CD19+ B cell repletion, defined as the time taken to reach baseline levels or lower limit of normal. (C) The mean absolute number of peripheral blood CD4+ or CD8+ T cells and CD56+ natural killer cells assessed by flow cytometry. (D) The adjusted (by geographical region) annualized relapse rate mean during the trial 0-24wk, the trial extension period between week 24-96, the whole treatment-free follow-up between week 96-144wk and the B cell repletion study during week 120-144, (E) The mean number of T1 gadolinium-enhancing lesions detected by magnetic resonance imaging during the placebo-controlled trial period for placebo (n=54), IFNβ (n=52) and 600mg ocrelizumab (n = 51) and for the treatment-free safety follow-up between week 96-120 (n=36) following 4 cycles of ocrelizumab (F) The proportion of people with 6 month confirmed disease progression assessed by worsening of the Expanded Disability Status Scale based on the intention to treat population. The results are a facsimile of those presented previously (20, 21).

The phase II trial demonstrated a significant (p=0.0019) reduction in the adjusted annualised relapse rate between placebo and ocrelizumab treatment from 0.557 (n=54) to 0.127 (n=55) at the 48-week analysis period after one treatment cycle (**Table 1, Figure 1D**. (2)). In addition to a relatively quiescent clinical picture, magnetic resonance imaging of the brain revealed no new T1 gadolinium-enhancing (**Figure 1E**) or T2 lesions in 36 pwMS having undergone imaging (20,21). The proportion of pwMS with confirmed 6-month disability progression remained low in all groups treated with ocrelizumab (20,21) (**Figure 1F**). Although treatment was not infallible and relapses occurred, efficacy appeared to be maintained through 3-4 treatment cycles of treatment of the extension study and the year-long safety and B cell monitoring period (**Figure 1D. Table 1**). Therefore, 3-4 cycles of 600mg ocrelizumab seems to induce disease inhibition that is durable during an 18-month treatment-free period, suggesting a long-term benefit from a short-term treatment cycle.

The safety issues related to 2 years of ocrelizumab treatment have been addressed within the phase III clinical trial programme involving 827 people with relapsing MS randomised to receive six-monthly 600mg ocrelizumab dosing (3). However, within the phase II study cohort, there were 174 adverse events, 11 serious adverse events (2 of which led to study withdrawal), 89 infections and 4 serious infection events within the three 600mg ocrelizumab extension study arms (n=151) during the first year of ocrelizumab treatment (**Table 2** (20)). This compared to only 72 adverse events, 3 serious events (salivary duct inflammation and a fatal injury 14 months after treatment due to a work-related trauma event deemed unrelated to treatment), 34 infection events and no serious infection events within the year of treatment-free follow-up. This suggested that there were more adverse events and infections during the first 24 weeks of treatment than during the last 24 weeks of the treatment free period (**Table 2**). However, some individuals are likely to be more susceptible to adverse events and may drive up AE frequency early on. Importantly, there were fewer pwMS completing the 120-week study (n=130) than entered the extension study and therefore those remaining within the study may be selected for responding well to treatment. This could account for the magnitude of reduced adverse events during the treatment-free period, but suggests potential benefit from avoiding drug treatment (**Table 2**). Perhaps adverse events should have increased as hypogammaglobulinemia and increased susceptibility to infection normally may develop over time. Whilst these effects may have been limited by the influence of a “drug holiday”, these elements will only be elucidated through larger follow-up studies.

**Table 2.** Reduced infections during the drug-free treatment period in the ocrelizumab phase II extension trial

| Treatment | Analysis | Serious/Adverse Events |  | Serious/Infection |  |
| --- | --- | --- | --- | --- | --- |
|  |  | 3 dosing cycles | 4 dosing cycles | 3 dosing cycles | 4 dosing cycles |
| Ocrelizumab | First 24 weeks | 1.9%/71.7% | 1.8%/63.6% | 1.9%/34.0% | 0.0%/43.6% |
| Ocrelizumab | Second 24 weeks | 2.0%/50.0% | 1.8%/54.0% | 0.0%/12.0% | 2.0%/36.0% |
| Treatment-Free | Second to Last 24 weeks | 0.0%/32.7% | 0.0%/32.7% | 0.0%/16.3% | 0.0%/18.8% |
| Treatment-Free | Last 24 weeks | 2.2%/26.1% | 2.2%/26.1% | 0.0%/10.9% | 0.0%/6.5% |

Data relating to the frequency of adverse events/serious adverse events and infections/serious infections following treatment with four six-monthly cycles of 600mg ocrelizumab (week 0-96) or after an infusion of placebo (week 0-24) followed by three cycles ocrelizumab. This was followed by an 18-month treatment free period (week 96-144). Results from the last 24 weeks of the study represent weeks 120-144 of the open label extension study. The beta interferon group was not analysed to avoid any influence of immunomodulation by the treatment (20).

## DISCUSSION

This study suggests that ocrelizumab may be a selective immune-reconstitution therapy, with a long-term efficacy from a short-term treatment cycle. As such, it was found that 12-18 months after the last infusion of 3 cycles of 600mg ocrelizumab, the levels of disease activity appear to be similar to that seen in the phase III extension studies following 6 cycles of ocrelizumab (20-22). However, caution is needed when comparing different studies and given the low numbers of people involved in this study. However, this sustained treatment-effect comes with the apparent benefit of a reduced risk of serious infection, whilst being treatment-free. There appeared to be fewer adverse events in the 6 months at the end of the treatment-free period (week 120-144) compared to the first six months and is unsurprising as infusion reactions are a common adverse events (2). This occurs in about 34% (n=283/825) of people with relapsing MS and probably relates to the lytic cell syndrome that occurs due to antibody-mediated killing of cells (27). The frequency of these reactions are reduced by the second cycle of antibody administration, although still present in about half (n=106/227) of the people demonstrating infusion reactions on first cycle (27). This is perhaps consistent with the finding that many people will have fewer circulating B cells when retreated. However adverse events still remain higher than during the treatment-free period suggesting, with caveats, that it may be possible to reduce the risk:benefit balance by reduced dosing frequency.

The phase I extension study of rituximab (2000mg/cycle) in MS also reported maintained benefit 12 months after the last infusion (1). Similarly, a single dosing cycle of 1000mg rituximab followed by maintenance regime of daily glatiramer acetate showed treatment failure (>2 new lesions, relapse or accumulated disability) in 10/27 (37%) pwMS, with a median time to failure of 23 months (lower 95% confidence limit 14.6 months) in a 36-month follow-up (28). It remains to be seen if this would have any additional benefit over rituximab monotherapy. In addition, off-label studies with rituximab where treatment was halted demonstrated long-acting benefit and an absence of rebound disease activity phenomena after stopping therapy (29). It is currently not clear whether ocrelizumab will have a longer treatment response, as it has more effective depletion characteristics than rituximab (1,25,30). As such, it was found that 20% people began to repopulate by 6 months (30), whereas only 5% pwMS began to repopulate within 6 months after ocrelizumab and CD19 repopulation took longer than 1 year (23,25). Importantly, this repopulation will be driven by immature/transitional and mature B cells emanating from the bone marrow, as a stereotyped B cell repletion characteristic (1,11). The memory B cell compartment that may harbour the key pathogenic cells, will likely remain depleted for very much longer, as shown following a single rituximab administration (7, 11).

The influence of ocrelizumab on individual B cell subsets during the trials is currently unreported, but it maintains all CD19 B cell subsets in a nadir state (3) and its effects on B cell subsets is consistent with the type of response reported following rituximab treatment (7,9). Although the longevity of the treatment response beyond repopulation of total CD19+ cells shown here, is consistent with other B memory cell targeting, immune-reconstitution therapies in MS (5,6,12), it remains to be seen whether therapy of relapsing MS actually relates to depletion of memory B cells, as appears to occur in some other CD20-responsive autoimmunities (4,9,11). However, it is known that rituximab can deplete memory B cells for over 12 months in MS and that a substantial depletion is still evident 2-3 years or more in other CD20-responsive autoimmune diseases (7,11). This type of slow repopulation, as seen in the blood, may relate to the durable, yet reversible disease control following a single infusion cycle (28). However, it should be remembered that relapses can develop in the apparent absence of peripheral blood memory B cells, indicating that an important B cell compartment is elsewhere (11,30). Further study is required to determine whether and when disease breakthrough occurs after a limited number of cycles of ocrelizumab, as found with other continuously-delivered immunotherapies or whether long-term disease control is seen, as found in many people treated with current T and B cell targeting, immune-reconstituting therapies (12-14).

While response to therapy is more consistent with a B cell-directed mechanism of action and the hypothesis for the importance of targeting memory B cells to control relapsing MS (3,4,11). However, to support a T cell-centric view of MS pathogenesis, it has been suggested that the activity of rituximab and ocrelizumab could be related to the depletion of CD20+ T cells (7,8,31). However, the influence of ocrelizumab on T cell numbers, as shown here, and during the phase III studies CD4+ cells decreased by 0.8% and CD8+ T cells decreased by 8.6% after 96 week of treatment in relapsing MS, was relatively marginal (21,22,26). This suggests that it is unlikely to account for the high efficacy of ocrelizumab. This is further indicated by the fact that marked CD4 T cell depletion had limited influence on relapsing MS (32) and importantly that, whilst T cells do not express significant levels of CD19 message or protein, CD19-B cell depleting antibodies also reduce the formation of active MS lesions in a manner similar to CD20-depleting antibodies (31,33). Although a relative increase in T regulatory cells may contribute to efficacy (8), the slow increase may not correspond well to the rapid clinical response (8, 10). This demonstrates an activity that is perhaps not likely to be dependent on direct T cell targeting. Likewise, the suggestion that the activity of ocrelizumab is by blocking the formation of ectopic B cell follicles, is also unlikely to account for the activity of ocrelizumab in relapsing MS, given CD20 is not expressed by plasmablasts and plasma cells, and antibodies are largely (99.9%) excluded from the central nervous system (11,34). As such, B cells and oligoclonal immunoglobulin bands in the cerebrospinal fluid persist, at least after rituximab administration (11, 35). However, increasing exposure may be associated with limiting the accumulation of progressive disability, possibly indicating that CNS penetration of antibody could contribute to efficacy (36). However, B cells may act as essential antigen presenting cells that activate pathogenic T cells or they may target oligodendrocytes or nerves to participate directly in the pathogenesis of MS (4,8).

If there is a durable activity of CD20-depletion, as seen with other immune-reconstitution therapies (12,13), ocrelizumab would have enhanced utility in the management of MS because of its relatively lower side-effect profile and limited monitoring requirements, compared to other high-efficacy treatments (14). This could also lead to cost-effectiveness improvements that may help improve access to treatment, in particular to the primary progressive MS indication, where concerns about the cost-effectiveness has led to limitations to treatment (37). However, even if ocrelizumab is simply an agent that requires repeated treatments, lower frequency administration will have benefits for pwMS as it may help avoid serious infectious complications (15,16). Studies with rituximab in MS and other autoimmune conditions have indicated that through B cell monitoring, it may be possible to extend dosing intervals without loss of efficacy (9,11,38). Further studies are required to determine whether adaptive ocrelizumab dosing based on B cell counts can be of value in MS. Importantly, an extended 12-18 month, or potentially longer, treatment-free period could provide sufficient time for a drug-free pregnancy whilst under ongoing protection from disease activity. Ocrelizumab has an elimination half-life of about 26 days (23-25) meaning the CD20-depletion potential probably remains for many months after infusion (2 x 300mg dose) as less than a thirtieth of the clinical dose remains potently B cell depleting (39). B cells form within foetal liver by 9 weeks and circulate from 12 weeks from conception (40). Therefore, transplacental passage of antibody can lead to foetal and neonatal B depletion (41). To avoid this issue, effective contraception is advised for 6-12 months, depending on region, after the last dose of ocrelizumab (23,24). Whilst CD20-depletion of pregnant females and foetal B cell depletion has been tolerated and neonatal depletion is transient after birth, post-partum maternal relapses have been documented following B cell depletion (23,24,41). Therefore, further studies to confirm the safety, the optimal number of treatment-cycles and the longevity of disease inhibition during pregnancy are required to determine whether acceptable levels of disease control are possible and whether there is a similar or longer duration of activity than that found with rituximab treatment (1,28). Although ocrelizumab is humanised to potentially reduce the frequency of binding (0.4% in 96-week phase III trial N=3/807 (3) and neutralizing antibodies (0.1% in phase III trial. N=1/807 (3)) seen with chimeric antibodies (28.6% at week 48 in the phase I rituximab trial (1), these are also controlled by dose (39). As such, 20mg (2 x 10mg) ocrelizumab infusion induced ocrelizumab-specific antibodies in about 19% (n=7/36 to 72 weeks) of people compared to 0% (n=0/40 per group) of people treated with 2 x 200mg or 2 x 500mg ocrelizumab (39). Therefore, it would be important to ensure that reduced-dosing does not allow neutralizing antibodies to develop as antibody levels taper and immature B cells repopulate, which could prevent activity.

The data presented here also suggests that head to head studies of limited-dosing ocrelizumab, versus current standard, repeated-dosing of ocrelizumab, and importantly versus an established immune-reconstitution therapy are warranted. This would determine whether there is safety and cost-effectiveness benefit of extended dosing and importantly that there is indeed benefit, as the low numbers completing the studies may have skewed the positive effects. In addition, off-label use of other CD20 depleting antibodies may help inform on this issue. Alternatively, it may help to determine whether the additional innate immune cell or T cell depleting capacities, provides additional benefit that may contribute to long-term disease control (5,12).

## Data Availability

This is not applicable

